# Plasma β-amyloid in mild behavioural impairment – neuropsychiatric symptoms on the Alzheimer’s continuum

**DOI:** 10.1101/2021.01.09.21249145

**Authors:** Ruxin Miao, Hung-Yu Chen, Sascha Gill, James Naude, Eric E. Smith, Zahinoor Ismail, for the Alzheimer’s Disease Neuroimaging Initiative

## Abstract

**Introduction:** Simple markers are required to recognize older adults at higher risk for neurodegenerative disease. Mild behavioural impairment (MBI) and plasma β-amyloid (Aβ) have been independently implicated in the development of incident cognitive decline and dementia. Here we studied the associations between MBI and plasma Aβ_42_/Aβ_40_.

**Methods:** Participants with normal cognition (n = 86) or mild cognitive impairment (n = 53) were selected from the Alzheimer’s Disease Neuroimaging Initiative. MBI scores were derived from Neuropsychiatric Inventory items. Plasma Aβ_42_/Aβ_40_ ratios were assayed using mass spectrometry. Linear regressions were fitted to assess the association between MBI total score as well as MBI domain scores with plasma Aβ_42_/Aβ_40_.

**Results:** Lower plasma Aβ_42_/Aβ_40_ was associated with higher MBI total score (*p* = 0.04) and greater affective dysregulation (*p =* 0.04), but not with impaired drive/motivation (*p =* 0.095) or impulse dyscontrol (*p =* 0.29) MBI domains.

**Conclusion:** In persons with normal cognition or mild cognitive impairment, MBI was associated with low plasma Aβ_42_/Aβ_40_. Incorporating MBI into case detection can help capture preclinical and prodromal Alzheimer’s disease.

## Background

Alzheimer’s disease (AD) is a neurodegenerative condition characterized by abnormal β-amyloid (Aβ) and tau aggregation into plaques and tangles, resulting in cerebral dysfunction. ^1^ A clinical diagnosis of dementia is established by the presence of functional decline, at which point neuronal loss and AD pathology are extensive and likely irremediable. AD clinical trials have failed to identify disease-modifying treatments, hindered in part by poor recruitment and retention of early phase disease. ^2,3^ One unifying goal in dementia research is to recognize high-risk individuals in the preclinical and prodromal stages of AD, not only to provide timely intervention but to target recruitment for future research. Simple markers for risk are required. ^4^

Although cognitive changes and functional decline are the hallmarks of AD progression, non-cognitive markers are also associated with incident dementia. ^5^ Of these markers, neuropsychiatric symptoms (NPS) are simple and inexpensive to determine and can be captured at scale. Mild behavioural impairment (MBI) is a validated neurobehavioural syndrome characterized by the later-life emergence of persistent NPS ^6^ as an at-risk state for incident cognitive decline and dementia, and the initial manifestation of dementia for some. ^7–10^ While research has been reliant on detailed neuropsychological testing, CSF biomarkers, and PET imaging in the early detection of dementia, these markers are labour-intensive and expensive, which limits scalability. Plasma biomarkers could also satisfy the need for a cost-effective, minimally invasive approach to quantify early AD risk and to correlate with non-cognitive markers such as MBI as an initial screen.

A growing body of work has demonstrated the efficacy of plasma biomarkers in the evaluation of early neurodegeneration. ^11–13^ Plasma Aβ_42_/Aβ_40_ ratio is a promising AD biomarker that predicts cerebral amyloid and AD pathology in at-risk individuals. ^14^ Little research has explored the association between MBI and plasma biomarkers, ^15^ and to our knowledge, none with plasma amyloid. Here we assessed plasma Aβ associations with MBI. Informed by recent evidence on the association between MBI and amyloid PET, ^16^ we hypothesized that MBI symptomatology would be associated with lower plasma Aβ_42_/Aβ_40_, signifying increased amyloid burden in the brain.

## Methods

### Study Population

#### Alzheimer’s Disease Neuroimaging Initiative (ADNI)

Data used in the preparation of this article were obtained from the Alzheimer’s Disease Neuroimaging Initiative (ADNI) database (adni.loni.usc.edu). The ADNI was launched in 2003 as a public-private partnership, led by Principal Investigator Michael W. Weiner, MD. The primary goal of ADNI has been to test whether serial magnetic resonance imaging (MRI), positron emission tomography (PET), other biological markers, and clinical and neuropsychological assessment can be combined to measure the progression of mild cognitive impairment (MCI) and early Alzheimer’s disease (AD). ADNI is currently in its 4 ^th^ iteration at the time of writing. All data used in this study were accessed from ADNI before October 19 ^th^, 2020.

### Participants

Participants were between 55-90 years of age (inclusive) and spoke either English or Spanish. They were accompanied by study partners. Informed consent was obtained from all participants before study enrollment. We selected participants with MMSE scores between 24-30 (inclusive). Exclusion criteria in ADNI included concurrent neuropsychiatric diagnoses such as clinically significant depression, psychosis, or non-Alzheimer’s neurological disorders.

### Measures

#### Clinical variables

Age, sex, and education were included to assess for potential associations with plasma Aβ_42_/Aβ_40_. Cognitive status (normal cognition [NC] or mild cognitive impairment [MCI]) was determined at the initial visit based on ADNI criteria (https://adni.loni.usc.edu/wp-content/uploads/2010/09/ADNI_GeneralProceduresManual.pdf).

#### Neuropsychiatric variables

In ADNI, NPS were assessed using the Neuropsychiatric Inventory (NPI). ^17^ Based on a published algorithm, ^18^ NPI items were used to derive the five MBI domains: decreased drive/motivation (NPI apathy/indifference); emotional dysregulation (NPI depression, anxiety, elation/euphoria); impulse dyscontrol (NPI agitation/aggression, irritability, aberrant motor behaviour); social inappropriateness (NPI disinhibition); and abnormal thoughts/perception (NPI delusions, hallucination). For each domain, domain scores were calculated as the product of domain frequency (range of 0 to 4) and severity (range of 0 to 3). Scores across the five transformed MBI subdomains were added to generate a total MBI score (theoretical range of 0 to 60).

#### Biomarkers

In the ADNI database, analysis of plasma Aβ_42_ and Aβ_40_ was completed by the Bateman Laboratory. ^19^ Participants were given a stable isotope tracer containing L-[U-^13^C _6_] leucine, and target Aβ isoforms were immunoprecipitated from plasma via anti-Aβ mid-domain antibody.

After digestion with Lys-N protease, reconstituted digests were subjected to liquid chromatography-tandem mass spectrometry. ^19^ To determine plasma Aβ_42_/Aβ_40_, assays were conducted using the Thermo Scientific TSQ Altis Triple Quadrupole Mass Spectrometer.

### Sample

Participants with available plasma amyloid were selected. NC and MCI participants with NPI data and plasma Aβ_42_/Aβ_40_ measurements were included in the final analysis (n=139). A detailed flowchart of how the final sample was obtained is shown in **Figure 1**.

**Figure 1.**
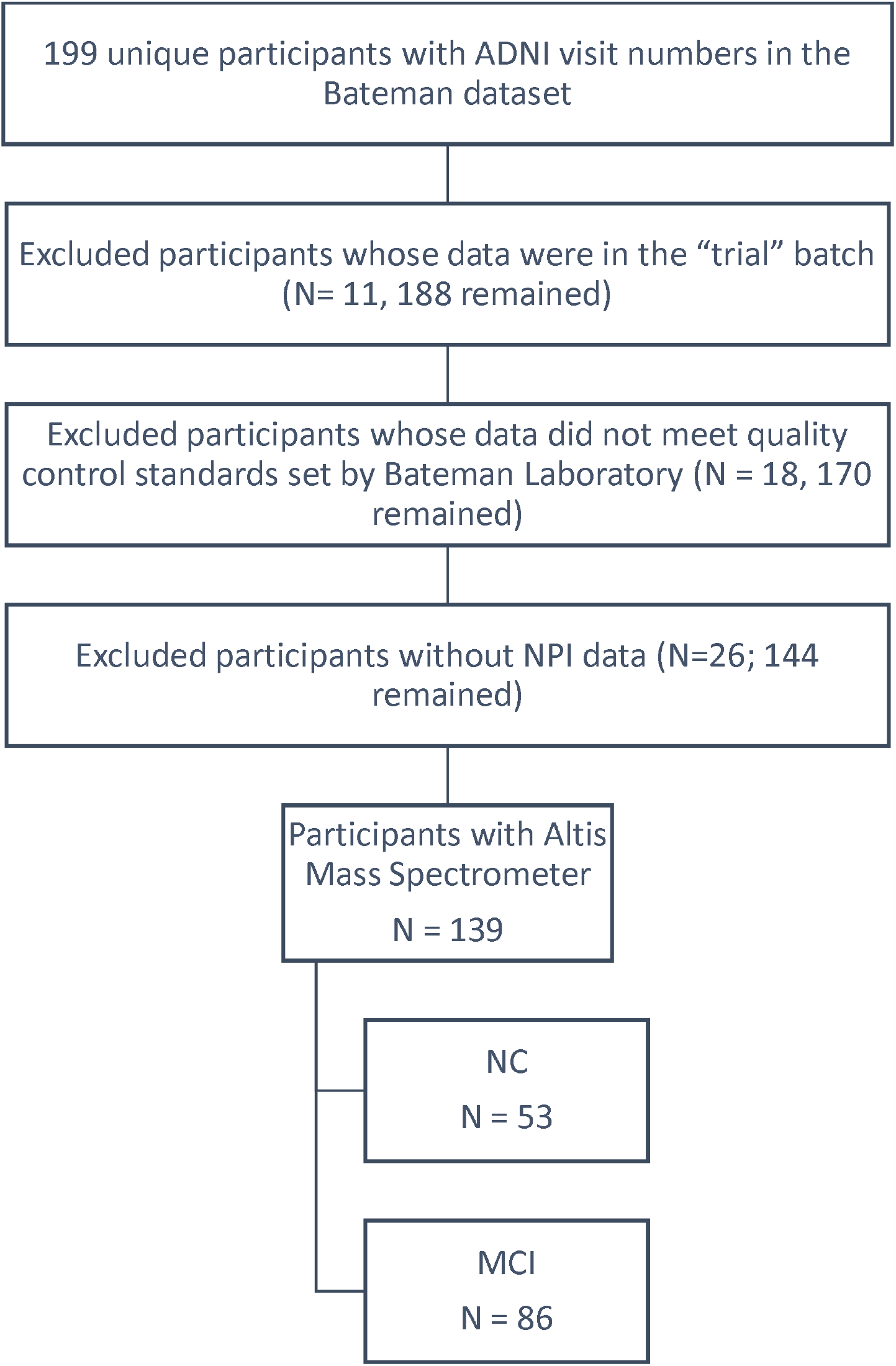
Flowchart showing how the sample populations were obtained from the ADNI cohort

### Statistical Analysis

Statistical analysis was performed using R (version 3.6.2). Linear regressions were fitted to assess the association between MBI total score as well as MBI domain scores with the outcome variable, plasma Aβ_42_/Aβ_40_. The domain scores included the following: affective dysregulation, drive/motivation, and impulse dyscontrol. Abnormal thoughts/perception and social inappropriateness were low frequency domains in this sample, precluding analysis. The models controlled for participants’ age, sex, education, and diagnostic status (NC or MCI).

## Results

Sample characteristics, including basic demographic information such as age, sex, and education, are shown in **Table 1**. A total of 139 participants were included in the analysis, with 86 in the NC group, and 53 in the MCI group.

**Table 1.** Sample Characteristics

| **Characteristics** | **Overall**<br>**N=139** | **NC**<br>**n=86** | **MCI**<br>**n=53** |
| --- | --- | --- | --- |
| Age (mean, SD) | 72.4 (7.6) | 73.9 (6.3) | 70.1 (6.5) |
| Female sex, (n, %) | 71 (51.8) | 45 (52.3) | 26 (49.1) |
| Education years, (median, Q1-Q3) | 17 (15-18) | 17 (15-18) | 17 (16-18) |
| MBI total score (mean, Q1-Q3) | 0.54 (0-1) | 0.41 (0-0) | 0.75 (0-1) |
| Any MBI symptoms (%) | 39 (28.1) | 20 (23.3) | 19 (35.8) |
| Affective dysregulation domain score (mean, Q1-Q3) | 0.22 (0-0) | 0.14 (0-0) | 0.36 (0-1) |
| Drive/motivation domain score (mean, Q1-Q3) | 0.058 (0-0) | 0.047 (0-0) | 0.075 (0-0) |
| Impulse dyscontrol domain score (mean, Q1-Q3) | 0.22 (0-0) | 0.20 (0-0) | 0.26 (0-0) |
| A$\beta_{42}$/A$\beta_{40}$ (mean, SD) | 0.123 (0.0133) | 0.122 (0.0131) | 0.124 (0.0136) |

Linear regressions revealed that a higher MBI total score was associated with decreased plasma Aβ_42_/Aβ_40_ (*p* = 0.04) (**Table 2**). An association with age was also observed such that older patients had lower Aβ_42_/Aβ_40_ (*p* = 0.003). All other variables, including sex, education, and baseline diagnosis of MCI versus NC, were not associated with plasma Aβ_42_/Aβ_40_. Further analyses of MBI domains showed a significant relationship between higher MBI affective dysregulation and lower plasma Aβ_42_/Aβ_40_ (*p =* 0.04) (**Table 3**). However, neither decreased motivation (**Table 3;** *p =* 0.095) nor higher impulse dyscontrol (**Table 3;** *p =* 0.29) were associated with lower plasma Aβ_42_/Aβ_40._

**Table 2.** The association between Mild Behavioural Impairment (MBI) and plasma Aβ_42_/Aβ_40_

|  | Unstandardized Coefficients |  |  |
| --- | --- | --- | --- |
|  | Estimate | Std. Error | Pr ( > \|t\| ) |
| (Intercept) | $1.58 \times 10^{-1}$ | $1.51 \times 10^{-2}$ | **<0.001** |
| Age (per year) | $-2.02 \times 10^{-3}$ | $9.68 \times 10^{-4}$ | **0.003** |
| Male sex | $3.42 \times 10^{-4}$ | $4.56 \times 10^{-4}$ | 0.328 |
| Education (per year) | $-2.23 \times 10^{-3}$ | $2.27 \times 10^{-3}$ | 0.455 |
| MCI versus NC | $1.58 \times 10^{-1}$ | $1.51 \times 10^{-2}$ | 0.478 |
| MBI total score (per additional point) | $-2.02 \times 10^{-3}$ | $9.68 \times 10^{-4}$ | **0.039** |

**Table 3.**
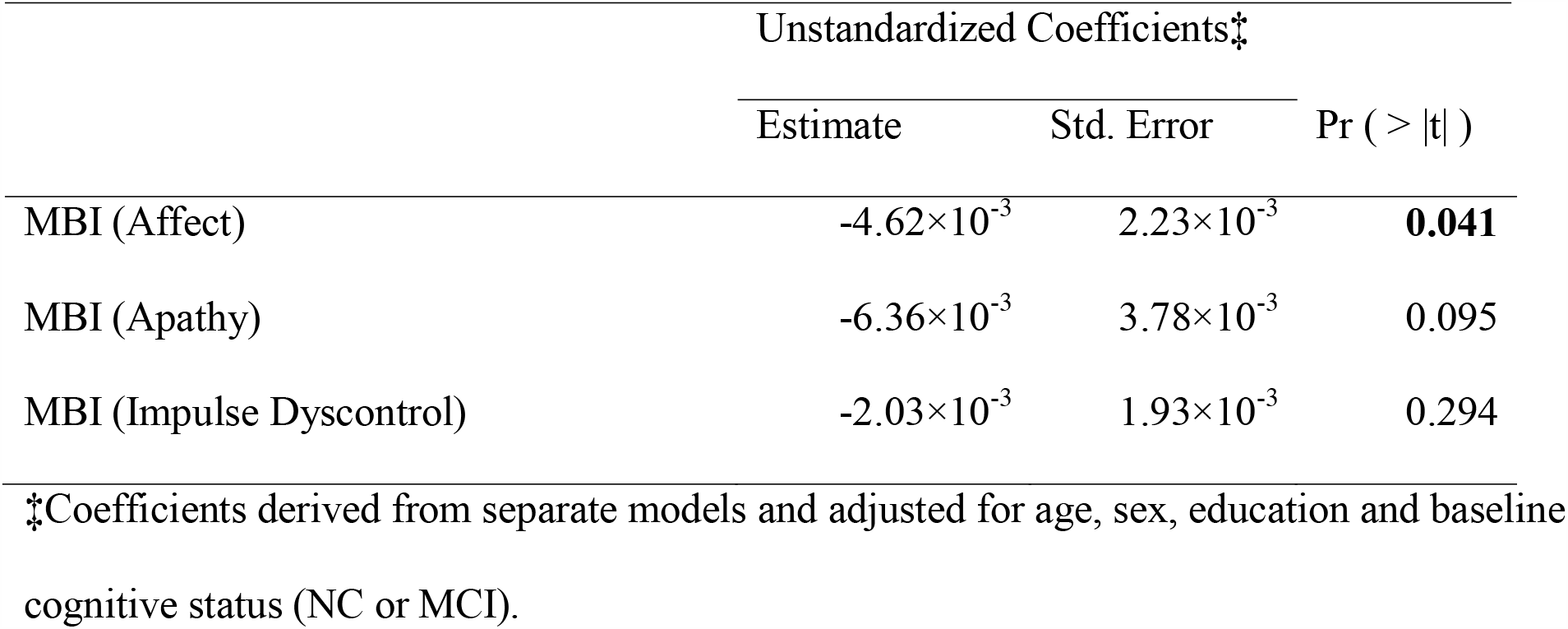
The association between Mild Behavioural Impairment subdomains and plasma Aβ_42_/Aβ_40_

## Discussion

In a sample of 139 participants with NC and MCI, those with greater MBI burden had significantly lower plasma Aβ_42_/Aβ_40_. Looking at MBI subdomains, decreased plasma Aβ_42_/Aβ_40_ was significantly associated with greater MBI affective dysregulation, but not MBI decreased motivation or impulse dyscontrol. To our knowledge, this study is the first to demonstrate the relationship between MBI and plasma Aβ_42_/Aβ_40_.

Amyloid plaques, particularly Aβ_42_, have been implicated in the pathogenesis of AD. ^1^ A large body of work has shown an inverse association between CSF Aβ_42_ levels and the degree of cerebral amyloid. ^1^ Plasma Aβ levels positively correlate with their CSF levels ^20^ and can be detected with high precision and specificity. ^19^ Plasma Aβ_42_/Aβ_40_ has shown promise in supporting the diagnosis of AD, ^21–23^ and has better diagnostic accuracy than plasma Aβ_42_ or Aβ_40_ alone. ^14,24^ Further, plasma Aβ_42_/Aβ_40_ has predictive value for abnormal amyloid density in the brain ^11–13^ and can work in conjunction with other plasma AD markers to detect brain atrophy and risk of clinical progression to dementia in NC and MCI populations. ^13^ Some early studies using older assays disputed the prognostic value of plasma Aβ_42_/Aβ_40_ for incident dementia, ^25^ while maintaining the consensus that plasma Aβ_42_ and Aβ_40_ concentrations increase significantly with age. ^1^ This consensus is consistent with our findings such that age was a significant covariable in the regression models. However, the predictive performance of Aβ_42_/Aβ_40_ is not influenced by age. ^14^ Newer research has identified high levels of plasma Aβ at baseline as a risk factor for future neurodegeneration in NC populations. These studies have also implicated decreasing plasma Aβ_42_ and Aβ_42_/Aβ_40_ in early AD progression. ^26,27^ Thus, a declining Aβ_42_/Aβ_40_ ratio may portend the development of clinical dementia symptoms. Differences in the association between plasma Aβ_42_/Aβ_40_ and incident dementia between older and newer studies may be explained by the improving specificity and accuracy of Aβ detection using mass spectrometry ^19^ compared to earlier methods such as enzyme-linked immunosorbent assay. ^25^ Overall, plasma Aβ_42_/Aβ_40_ does appear to be an appropriate biomarker for the screening of pre-dementia states.

NPS are present in both preclinical and prodromal disease. In our sample, MBI symptoms were present in 39/139 (28.1%) participants. In a population-based cohort, using a similar methodology for NPI score transformation as ours, NPS were present at rates of 30.8% in NC and 47.1% in subjective cognitive decline (SCD). ^28^ According to one systematic review, NPS are also common in prodromal AD at 35-85%. ^29^ The emergence of NPS in older adults can herald cognitive decline in both NC ^8^ and MCI. ^30^ In a study of 2769 National Alzheimer’s Coordinating Center participants with a Clinical Dementia Rating (CDR) of 0 at baseline, 23.1% with MBI developed cognitive and functional decline to a CDR > 0 at 3-year follow-up. ^8^ While the risk of progression from MCI to dementia is around 10-15% at baseline, ^31^ this incidence jumps to 25% in the context of NPS. ^32^ MBI provides a validated approach to NPS characterization in at-risk older adults. ^33^ Prevalence of MBI varies depending on the setting and approach to MBI case ascertainment. In a psychiatric outpatient clinic, MBI prevalence was as low as 3.5% ^9^ using chart review, whereas prevalence was higher at 5.8% in SCD ^34^ and 14.2% in MCI ^35^ in a primary care setting using the MBI checklist (MBI-C). ^33^ A sample of memory clinic patients had a high MBI prevalence of 76.5% in SCD and 85.3% in MCI, ^18^ determined using the NPI-Q. ^36^ Using the same methodology as the present study, population estimates of MBI varied according to cognitive status, from 43.1% in at-risk NC to 48.9% in MCI. ^28^ Overall, prevalence estimates using the MBI-C are lower than those using the NPI. ^34,35^ Nonetheless, MBI is associated with cognitive decline throughout the AD continuum, including faster progression in NC ^7^ and greater incidence of dementia. ^9,10^ MBI also confers a greater risk of progression to dementia than MCI without MBI. ^37^

MBI has several biological correlates, which the literature is just beginning to explore. Known associations exist between MBI and biological markers of AD, including Aβ-positron emission tomography (PET), ^16^ cerebrospinal fluid (CSF) tau and tau-PET, ^38,39^ white matter atrophy, ^40,41^grey matter atrophy, ^42,43^ plasma neurofilament light (NfL), ^15^ and AD risk gene loci. ^44,45^ The current study extends the literature on the biological basis of MBI. For some, MBI captures the neurobehavioural symptomatology of early phase disease as evidenced by its inverse relationship with plasma Aβ_42_/Aβ_40_. Our data corroborate the use of MBI as a sensitive instrument for tracking pre-dementia states. There were no significant differences in plasma Aβ_42_/Aβ_40_ between NC and MCI groups. This supports the utility of MBI in identifying at-risk individuals regardless of baseline cognitive status (NC or MCI), and suggests that for some individuals, MBI may be more sensitive to early Aβ changes than cognition. Importantly, in the NIA-AA clinical staging for AD, NPS are incorporated in a way that is aligned with MBI. Specifically, individuals can be placed in stage 2 AD based on neurobehavioural symptoms alone, in the absence of cognitive decline. These “mild neurobehavioural changes” should have a clearly defined recent onset, which persist and cannot be explained by life events alone. ^46^ Our data further support the NIA-AA clinical staging by linking neurobehavioural symptoms with Alzheimer’s proteinopathies.

Although the connection between Aβ_42_/Aβ_40_ and cognitive decline is well-established, relatively less is known about AD biomarkers and their role in the neuropsychiatric sequelae of disease. In this study, affective dysregulation was the only MBI domain significantly associated with decreased plasma Aβ_42_/Aβ_40_. MBI affective dysregulation consists of NPI depression, anxiety, and elation/euphoria, and relates to changes in regulating emotional tone as a consequence of the structural and functional changes associated with AD. ^6^ The association between AD biomarkers and anxiety is supported in the literature, with three studies showing a relationship between decreased CSF Aβ_42_ and anxiety. ^47–49^ The relationship between Aβ and depression is more uncertain. Some studies identified no relationship between decreased CSF Aβ_42_ and increased depression in samples of mixed cognitive status; ^47,48,50^ others revealed interactions with mood disturbance in NC ^49,51^ and chronic subsyndromal depressive symptoms in MCI. ^52^ Plasma Aβ levels have been associated with depression, such that higher baseline plasma Aβ_42_ was a predictor for first depressive episode in NC older adults, and progression to AD at 5 years. ^53^ Higher baseline Aβ_40_ was also associated with depressive symptoms in the context of dementia at 11-year follow-up. ^54^ In a longitudinal study of cognitively normal older adults with no mood symptoms or mild depression at baseline, worsening depressive symptoms over 2-7 years were significantly associated with cognitive decline, and this effect was moderated by the presence of cortical amyloid on PET imaging. ^55^ However, amyloid deposition was less severe in older adults with concurrent major depression than in older adults without depression. ^56^ In NC samples, it appears that the pathophysiology of dementia differs in MBI (as characterized by the later-life onset of persistent NPS) versus in pre-existing psychiatric conditions. Although the data are equivocal, if there is a true acceleration of cognitive decline in older adults with emergent depression and concurrent amyloid burden, treatment of depressive symptoms might offer an opportunity to delay the progression of dementia symptoms. ^55^

We found a trend toward an association between lower plasma Aβ_42_/Aβ_40_ and MBI decreased motivation. MBI decreased drive/motivation, as denoted by NPI apathy/indifference, may have a tenuous association with amyloid biomarkers in the literature. The relationship between lower CSF Aβ_42_ and apathy was seen in one study ^47^ but not in several others. ^48,49,57^ However, new work demonstrated a significant association between Aβ-PET and MBI decreased motivation. ^39^ As well, another study determined that new-onset apathy, agitation, and irritability predicted faster progression from MCI to AD. ^58^ A systematic review and meta-analysis determined that the most common NPS in AD is apathy (49%), followed by depression (42%) and aggression (40%). ^59^ Considering the high prevalence of MBI decreased motivation in dementia, it is unsurprising that the association between Aβ_42_/Aβ_40_ and apathy trended towards significance in this small sample.

Finally, subdomain analysis revealed no association between plasma Aβ_42_/Aβ_40_ and MBI impulse dyscontrol. There have been several studies related to the subdomain of MBI impulse dyscontrol, which is comprised of NPI agitation/aggression, irritability, and aberrant motor behaviour. One review found a consistent relationship between CSF AD biomarkers (Aβ_42_, t-tau, and p-tau) and agitation/aggression in a mixed population of MCI and AD, with no other NPS correlates. ^57^ In contrast, another review paper investigating CSF Aβ_42_ and agitation/aggression in AD did not demonstrate a clear association. ^60^ In summary, the association between Aβ_42_ and MBI impulse dyscontrol-related NPS is inconsistent and somewhat surprising given the natural history of impulse dyscontrol symptoms in the evolution of dementia. Several longitudinal studies have identified emergent impulse dyscontrol symptoms among the earliest behavioural changes associated with incident cognitive decline and dementia. ^31,61,62^ Two studies found that impulse dyscontrol symptoms were comparable to hippocampal atrophy for prognostication of cognitive decline and dementia, ^41,63^ associated with both grey and white matter changes in advance of dementia. ^40^ Thus, the link between AD biomarkers and MBI subdomains can be challenging to interpret, and further investigation is required to explore these relationships.

### Limitations and Future Directions

Our study has some limitations. MBI total score was extrapolated from composite scores on corresponding NPI items, thus approximating MBI. The MBI-C is the case ascertainment instrument developed to capture MBI in accordance with the International Society to Advance Alzheimer’s Research and Treatment - Alzheimer’s Association MBI criteria. ^33^ The MBI-C captures emergent and persistent NPS over a period of 6 months or greater, whereas NPI addresses a point estimate of NPS over a 1-month period. Despite the established algorithm of transformation of NPI items to MBI domains, point estimates of NPS are more susceptible to the inclusion of reactive or transient NPS, decreasing specificity and potentially inflating MBI prevalence. ^18,28^ With our current screening measures, it can be difficult to distinguish between chronic and recurrent psychiatric conditions in late life, versus new-onset neuropsychiatric symptoms in the context of neurodegeneration. Thus, ADNI exclusion criteria around pre-existing psychiatric illnesses may preclude the participation of some older adults with MBI if the pre-existing symptoms are of relatively recent onset. Therefore, our ADNI sample might not be fully representative of MBI. Finally, the NPI was developed for and validated in a clinical AD population, and though it has since been used extensively in MCI, its ability to evaluate NPS in NC is not as well defined.

To elucidate the relationship between MBI and plasma biomarkers of AD, future studies should more precisely measure NPS. The MBI-C would allow for a standardized approach to documenting NPS in pre-dementia populations, ^33^ which may increase the specificity and yield of NPS subdomain analyses. Unfortunately, our data for NPS were limited to a single time-point due to linkage with the plasma biomarker analyses. Previous studies have successfully used NPS captured at two time points to approximate the persistence criterion of MBI, ^8,63^ and subsequent studies could use this approach if MBI-C data are not available. It would be valuable to track the natural history of MBI as it relates to longitudinal changes in plasma Aβ_42_/Aβ_40_. There are limited studies on the effects between AD biomarkers and MBI psychosis and social cognition, and a larger sample size may better characterize these relatively rare NPS. Finally, our study suggests that MBI predicts early Aβ pathology regardless of baseline cognitive status (NC or MCI), and thus potentially in advance of substantial cognitive changes. This merits replication in future analyses.

## Conclusion

Our study provides evidence for the biological association between MBI and plasma Aβ_42_/Aβ_40_, a validated marker of brain beta-amyloid. ^27^ Therefore, MBI may have utility as an accessible case ascertainment approach in AD clinical trial recruitment. More research is required to delineate the patterns of association between AD biomarkers and MBI symptoms over time.

## Data Availability

Data used in the preparation of this article were obtained from the Alzheimer's Disease Neuroimaging Initiative (ADNI) database (adni.loni.usc.edu).

http://adni.loni.usc.edu/

## Acknowledgements

Data collection and sharing for this project was funded by the Alzheimer’s Disease Neuroimaging Initiative (ADNI) (National Institutes of Health Grant U01 AG024904) and DOD ADNI (Department of Defense award number W81XWH-12-2-0012). ADNI is funded by the National Institute on Aging, the National Institute of Biomedical Imaging and Bioengineering, and through generous contributions from the following: AbbVie, Alzheimer’s Association; Alzheimer’s Drug Discovery Foundation; Araclon Biotech; BioClinica, Inc.; Biogen; Bristol-Myers Squibb Company; CereSpir, Inc.; Cogstate; Eisai Inc.; Elan Pharmaceuticals, Inc.; Eli Lilly and Company; EuroImmun; F. Hoffmann-La Roche Ltd and its affiliated company Genentech, Inc.; Fujirebio; GE Healthcare; IXICO Ltd.; Janssen Alzheimer Immunotherapy Research & Development, LLC.; Johnson & Johnson Pharmaceutical Research & Development LLC.; Lumosity; Lundbeck; Merck & Co., Inc.; Meso Scale Diagnostics, LLC.; NeuroRx Research; Neurotrack Technologies; Novartis Pharmaceuticals Corporation; Pfizer Inc.; Piramal Imaging; Servier; Takeda Pharmaceutical Company; and Transition Therapeutics. The Canadian Institutes of Health Research is providing funds to support ADNI clinical sites in Canada. Private sector contributions are facilitated by the Foundation for the National Institutes of Health (www.fnih.org). The grantee organization is the Northern California Institute for Research and Education, and the study is coordinated by the Alzheimer’s Therapeutic Research Institute at the University of Southern California. ADNI data are disseminated by the Laboratory for Neuro Imaging at the University of Southern California.

## Declaration of Conflicting Interests

The author(s) declared no potential conflicts of interest with respect to the research, authorship, and/or publication of this article.

## Funding

The author(s) received no financial support for the research, authorship, and/or publication of this article.

## Notes

### Competing Interest Statement

The authors have declared no competing interest.

### Author Declarations

Approval to access and use the present data was obtained from the ADNI database. Data collection for this study was conducted according to Good Clinical Practice guidelines, the Declaration of Helsinki, US 21CFR Part 50 - Protection of Human Subjects, and Part 56 - Institutional Review Boards, and pursuant to state and federal HIPAA regulations. Written informed consent for the study was obtained from all subjects and/or authorized representatives and study partners before protocol-specific procedures were carried out. Approval from an ethical standards committee to conduct this study was received at contributing ADNI sites.

